# Cardiac Health Assessment using a wearable device before and after TAVI

**DOI:** 10.1101/2023.03.22.23287604

**Authors:** Rob Eerdekens, Jo M. Zelis, Herman ter Horst, Caia Crooijmans, Marcel van ‘t Veer, Daniëlle C.J. Keulards, Marcus Kelm, Gareth Archer, Titus Kuehne, Guus R.G. Brueren, Inge Wijnbergen, Nils P. Johnson, Pim A.L. Tonino

## Abstract

**Background:** Due to the aging of the population, the prevalence of aortic valve stenosis will increase dramatically in upcoming years. Consequently Transcatheter Aortic Valve Implantation (TAVI) procedures will also expand worldwide. Optimal selection of patients who benefit with improved symptoms and prognosis is key since TAVI is not without risk. Currently we are not able to adequately predict functional outcome after TAVI. Quality of life measurement tools and traditional functional assessment tests do not always agree and can depend on factors unrelated to heart disease. Activity tracking using wearable devices might provide a more comprehensive assessment.

**Objectives:** Identify objective parameters from a wearable device (the Philips Health Watch) associated with improvement after TAVI for severe aortic stenosis.

**Methods and results:** 100 patients undergoing routine TAVI wore a Philips Health Watch for one week before and after the procedure. Watch data were analyzed offline: 97 before and 75 after TAVI. Parameters like the total number of steps and activity time did not change, in contrast to improvements in the six-minute walking test (6MWT) and physical limitation domain of a questionnaire (transformed WHOQOL-BREF).

**Conclusions:** These findings in an elderly TAVI population show that watch-based parameters like the number of steps do not change after TAVI, unlike traditional 6MWT and QoL assessments that do improve. Basic wearable device parameters might be less appropriate for measurement of treatment effects from TAVI.

## Introduction

As transcatheter aortic valve implantation (TAVI) for severe aortic stenosis is increasingly used in the elderly population, including a high percentage of patients with substantial co-morbidity, improvement in quality of life is just as important as extending life expectancy.^1, 2^ Not all TAVI patients benefit from improved physical activity as assessed by a 6-minute walking test (6MWT) or quality of life (QoL) questionnaire.^3, 4^ However, these tests could be influenced by other factors and comorbidities like peripheral vascular disease for the 6MWT or depression for the QoL questionnaire. Another concern with such tools is that they merely provide a snapshot of a patient’s life, and might change under different circumstances. Consequently, an unbiased and longer-term tool to anticipate the benefit from TAVI would allow physicians and patients to personalize treatment and expectations.

In recent years digital health has begun to transform the medical world.^5^ Smart phones and health watches, in particular, have found their way into the clinic.^6^ These devices can detect atrial fibrillation^7^ and predict the wearer’s 5-year risk of dying.^8^ The wearable device used in this study, the Philips Health Watch,^9^ continuously measures physical parameters like heart rate, number of steps, and amount of physical activity. Combining parameters from the health watch might provide a more physiologic and comprehensive assessment of functional status before and after TAVI. After intervention for aortic stenosis, patient symptoms often improve, but do they objectively become more active as measured by a wearable tracker?

In this study, we evaluated the change in parameters collected by the Philips Health Watch in patients before and after TAVI in comparison to standard clinical and research tests (6MWT, quality of life questionnaire). We hypothesized that after a TAVI procedure, physiologic parameters such as step count, total activity time, and daily total energy expenditure would increase, whereas respiration rate and heart rate would decrease.

## METHODS

This prospective, exploratory study sought to identify parameters from the Philips Health Watch (DL8791, Philips, Stamford, CT, USA) that changed after successful TAVI. The study was performed in compliance with the Declaration of Helsinki and local regulations. All subjects gave written informed consent as approved by an independent medical ethics committee (MEC-U approval ID:W16.141).

### Study population

Between July 2017 and September 2018, 100 consecutive patients (≥18 years) were included with severe aortic valve stenosis undergoing a clinically-indicated TAVI after Heart Team decision. Exclusion criteria were immobility and not being able to wear an electronic health watch. All patients were recruited at the Catharina Hospital in Eindhoven.

### Study protocol

Before TAVI all patients underwent transthoracic echocardiography, computed tomography for valve sizing and access site evaluation, laboratory testing, and clinical assessment as per local protocol. Patients were screened at the outpatient clinic; eligible and consenting patients received the Philips Health Watch. The watch was placed around the patient’s wrist after configuration with patient-specific parameters (height, weight, resting heart rate, and birth year). It was locked in the time screen, thereby blinding patients from all activity parameters, and worn for a week before being returned for data extraction. TAVI took place within 3 to 6 months of the baseline assessment. Three months after the TAVI procedure, patients visited the outpatient clinic for follow-up and again wore the Philips Health Watch for one week. At baseline and follow-up a 6MWT and questionnaire (transformed WHOQOL-BREF) were administered.^10^

### Analysis of the health watch data

The Philips Health Watch is a wrist-worn, photoplethysmography based, heart rate and activity monitor (Figure 1). Once per minute it measures parameters like heart rate, respiration rate, step count, and total energy expenditure (number of calories needed to carry out physiologic functions like breathing and physical activity, but excluding the energy required for digesting food) as described previously by Hendrikx et al.^9^ Parameters are measured at a 1-Hz sampling rate and stored on the device as 1-minute average values. Data can be extracted via Bluetooth by means of an iPod, using a proprietary iOS application from Philips, and sent to a Philips Research server for use in analyses.

**Figure 1.**
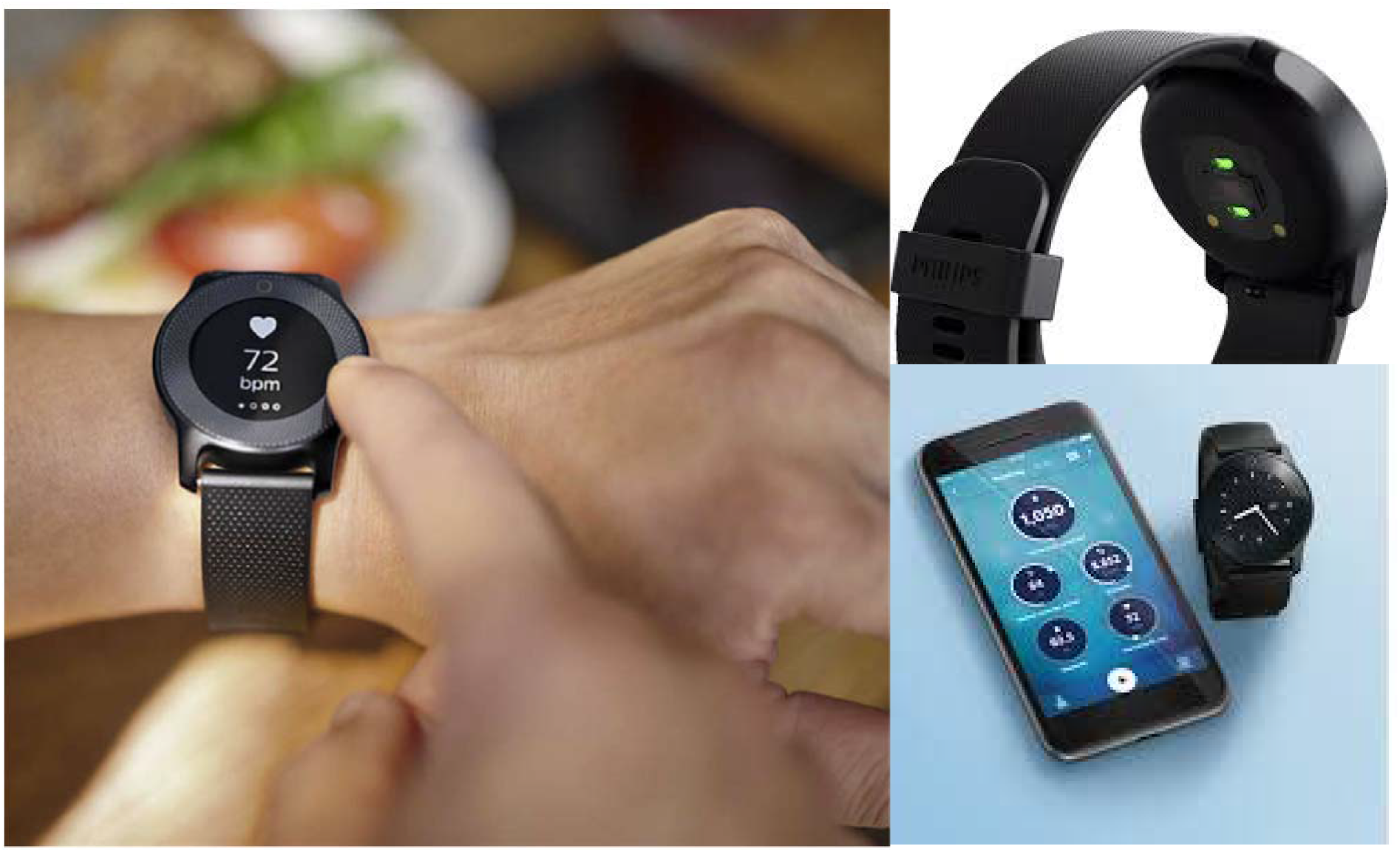
Philips Health Watch. The Philips Health Watch is a wrist-worn, photoplethysmography based, heart rate and activity monitor. It measures parameters like heart rate, respiratory rate, step count, total energy expenditure, and activity time. Measurements use a 1-Hz sampling rate and are stored on the device as 1-minute average values. Data can be extracted in the hospital via Bluetooth using the iPad application Watch Control and sent over Wi-Fi to the Philips Research server for analysis.

A full report including primary data from the watch and derived parameters consists of a summary averaged over one day (Table 3), distributions of heart rate and respiration rate (Figure 2), and log plot of the heart rate and total energy expenditure (Figure 3). Total energy expenditure is divided into subcategories of the Metabolic Equivalent Task (MET) scale. As the elderly TAVI population of this cohort seemed rather inactive, a subdivision of the MET scale was designed: basal activity corresponded to a MET of 1.5 to 2, light activity from 2 to 3, moderate activity from 3 to 6, and high activity from 6 upwards (we used standard thresholds for the last two categories).^11^

**Figure 2.**
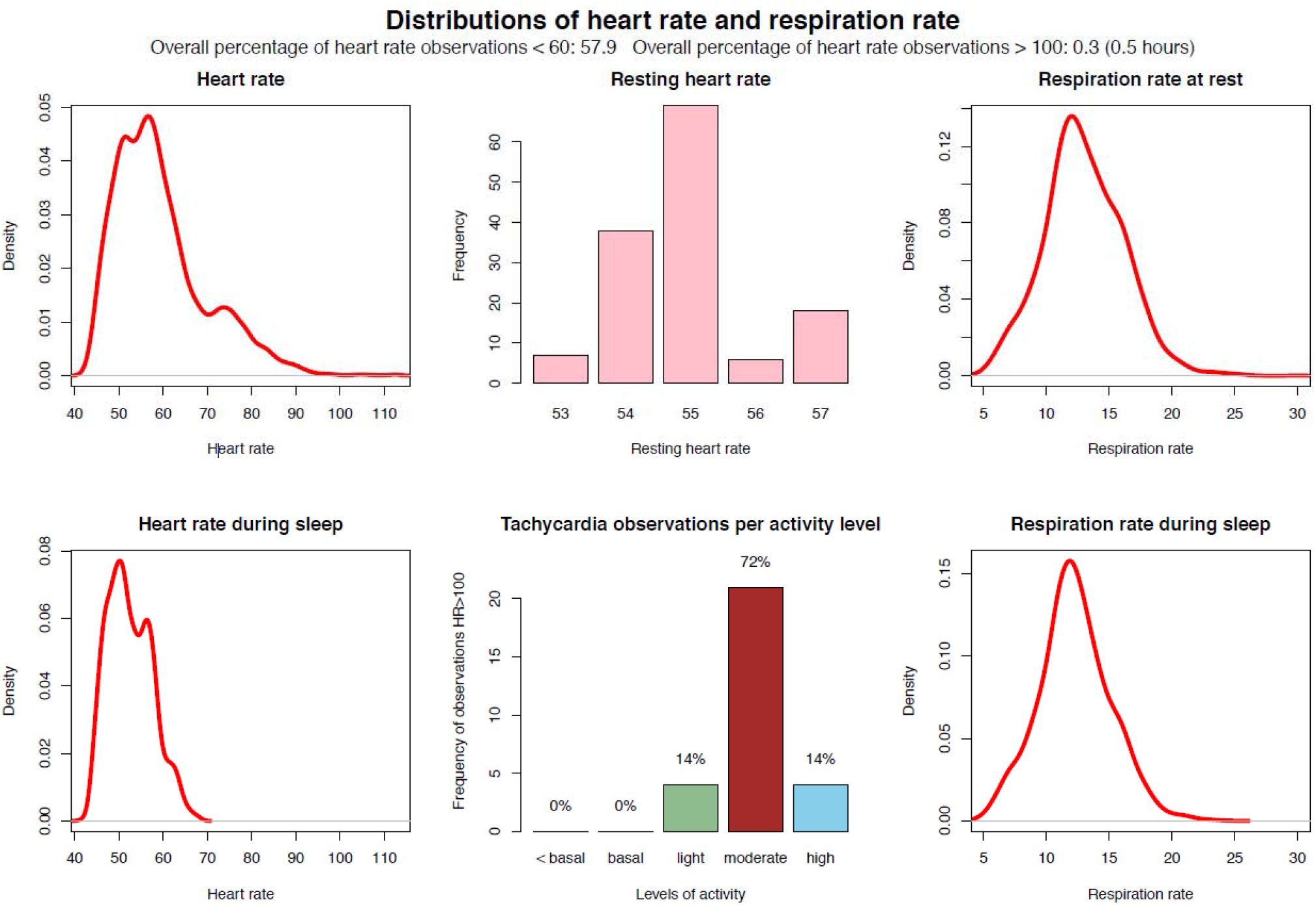
Distribution of heart rate and respiratory rate. This page of the output report from the health watch depicts density plots of heart rate and respiration rate during the day and during sleep. It also shows the frequency of resting heart rate and the distribution of activity levels for heart rate observations > 100 beats per minute.

**Figure 3.**
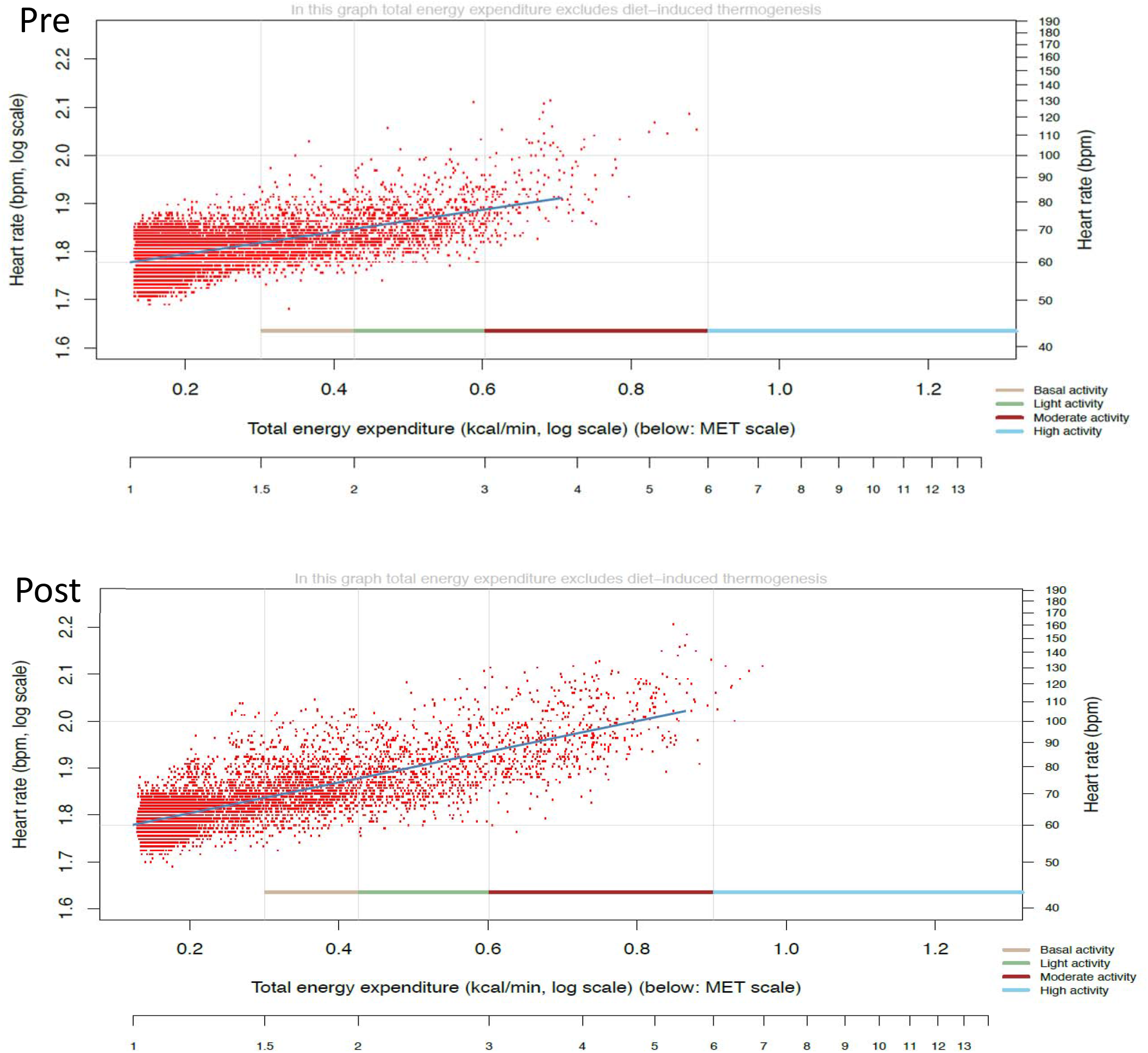
Log plot of heart rate vs. total energy expenditure. Another page of the report plots each heart rate and corresponding total energy expenditure on a log plot, divided into subcategories of the Metabolic Equivalent Task (MET) scale. The fitted line relates to the energy efficiency of the cardiovascular system, as detailed in the text.

Each red dot in Figure 3 represents a particular measurement: the 1-minute average heart rate and corresponding energy expenditure level. The fitted line quantifies the cardiac energy expenditure slope (CEES): as heart rate rises, more energy is needed to maintain the resulting hemodynamic state. When the slope is less steep, more energy is needed to maintain a heart rate of, for example, 60/minute. Conversely, when the slope is steeper, less energy is needed to maintain the same hemodynamic state. Potentially the steepness of the slope (CEES) serves as an indicator for the energy efficiency of the cardiovascular system.

The report and concept of CEES was proposed and made available as data derived from the raw data from the health watch by (the author from) Philips Research Eindhoven, and used in clinical data analysis at Catharina Hospital in Eindhoven.

### Statistical methods

Analyses were performed using SPSS Statistics software version 29.0 (IBM Corp). The data are displayed as mean values with standard deviations (SD) unless stated otherwise. Dichotomous variables are displayed as percentages (%) and absolute numbers (n). Applicable tests were two-tailed, and p<0.05 was considered statistically significant. Students T-tests were used when the compared variables had a normal distribution. Categorical variables were analyzed using the Chi-square test, Fisher’s exact test, or McNemar-Bowker test, whichever was appropriate. As this was an exploratory study, no sample size was pre-specified. Analyses were performed on the overall population (‘overall cohort’), men versus women (‘gender cohort’), above and under age 81 years (‘81 years cohort’), above and under age 85 years (‘85 years cohort’), and a cohort that had an increase in the 6MWT after TAVI of more than 40 meters (‘good responders cohort’).

## RESULTS

A total of 100 subjects were enrolled into the study. Their demographics and medications (before and after TAVI) are displayed in Table 1 and Table 2, respectively. After TAVI, 11 patients died and 14 patients were lost to follow-up. Data extraction for 3 patients failed before TAVI. Complete watch data were thus obtained in 97 (pre TAVI) versus 75 (post TAVI) patients. The population consisted of more men (57%) than women. Demographic characteristics were representative of a clinical TAVI population with a median age of 81.0 years, NYHA class II or higher in 92%, hypertension in 66%, and dyslipidemia in two-thirds of patients. All patients fulfilled guideline criteria for severe aortic stenosis. Operative characteristics can be found in the supplemental material (*Procedural data)*.

**Table 1.** Demographics at baseline

| **<u>Characteristic</u>** | **<u>Summary (n=100)</u>** | **<u>Good responders preTAVI (n=43)</u>** |  |
| --- | --- | --- | --- |
| Age (years) | 81.0 [76.0-84.0] | 81.0 [74.0-84.0] |  |
| Male | 57% | 51% |  |
| **<u>Risk factors</u>** |  |  |  |
| Active smoking | 8% | 12% | 0.19 |
| Hypertension | 66% | 72% | 0.15 |
| Dyslipidemia | 67% | 65% | 0.36 |
| Diabetes mellitus | 23% | 14% | 0.13 |
| **<u>Cardiac history</u>** |  |  |  |
| Prior myocardial infarction | 31% | 33% | 0.86 |
| Prior PCI | 41% | 49% | 0.33 |
| Prior CABG | 22% | 28% | 0.55 |
| **<u>Cardiovascular disease</u>** |  |  |  |
| Cerebral vascular disease | 18% | 14% | 0.37 |
| Peripheral vascular disease | 16% | 12% | 0.74 |
| COPD | 27% | 30% | 0.42 |
| Permanent pacemaker | 9% | 9% | 1.00 |
| **<u>Laboratory values</u>** |  |  |  |
| Hemoglobin (mmol/L) | 8.0 ± 0.9 | 7.9 ± 0.7 | 0.42 |
| hs-cTnT (ng/L) | 21.0 [14.0-37.8] | 23.5 [15.0-34.7] | 0.63 |
| NT-proBNP (pmol/L) | 1484 [835-3178] | 1250 [835-2763] | 0.79 |
| Creatinine (mg/dL) | 97.0 [77.0-119.0] | 101.0 [81.0-118.0] | 0.93 |
| **<u>Echocardiographic parameters</u>** |  |  |  |
| Left ventricular ejection fraction (%) | 56 [46-63] | 56 [42-64] | 0.62 |
| AV max velocity (cm/sec) | 424 [381-467] | 412 [386-464] | 0.96 |
| AV mean pressure gradient (mmHg) | 45 ± 14 | 42 ± 13 | 0.97 |
| AVA (cm<sup>2</sup>) | 0.8 ± 0.2 | 0.7 ± 0.1 | 0.70 |
| **<u>Symptoms</u>** |  |  |  |
| NYHA heart failure |  |  | 0.15 |
| class I | 8% | 9% |  |
| class II | 26% | 14% |  |
| class III | 57% | 63% |  |
| class IV | 6% | 12% |  |
| CCS angina ≥ III | 21% | 21% | 0.41 |
| Syncope | 9% | 7% | 0.42 |

Abbreviations: PCI = percutaneous coronary intervention; CABG = coronary artery bypass grafting; COPD = chronic obstructive pulmonary disease; hs-cTnT = high sensitive cardiac troponin T; NT-proBNP = N-terminal B-type natriuretic peptide; NYHA = New York Heart Association; AV = Aortic Valve; AVA = Aortic Valve Area; CCS = Canadian Cardiovascular Society grading of angina pectoris. Summary values represent number = % given 100 subjects. mean ± standard deviation or median [IQR].

**Table 2.** Medications

|  | Pre (N=80) (%) | Post (N=80) (%) | p value |
| --- | --- | --- | --- |
| Aspirin | 45 (56%) | 45 (56%) | 0.21 |
| Antiplatelet | 28 (35%) | 72 (90%) | < 0.001 * |
| Anticoagulation | 38 (47%) | 34 (43%) | 0.77 |
| Beta blocker | 56 (70%) | 38 (48%) | 0.26 |
| RAAS | 58 (73%) | 40 (50%) | 0.21 |
| Potassium sparing diuretic | 12 (15%) | 8 (10%) | 1.00 |
| Statin | 61 (76%) | 41 (51%) | 0.07 |
| Calcium channel blocker | 23 (29%) | 15 (19%) | 1.00 |
| Nitrates | 22 (28%) | 14 (18%) | 0.15 |
| Alpha blocker | 10 (13%) | 7 (9%) | 1.00 |
| Anti-arrhythmic | 5 (6%) | 5 (6%) | 1.00 |
| Insulin | 12 (15%) | 7 (9%) | 0.13 |
| Oral diabetic | 23 (29%) | 17 (21%) | 0.63 |
| Loop diuretic | 38 (48%) | 30 (38%) | 0.75 |

Abbreviations: RAAS: Renin-angiotensin-aldosterone-system. \* p value < 0.05

### Health Watch Parameters

Before TAVI versus after TAVI changes for all health watch parameters are displayed in Table 3 and Table S3 for the good responders cohort. Notably, in the total cohort no parameter changed significantly. For the female cohort, there was a small increase in the light-to-moderate activity time (206.6 before vs 207.3 minutes after TAVI, p = 0.03). For the below 81 years and good responders cohort, there was an increase in daily moderate activity: 14.2 versus 39.3 minutes (p = 0.02) and 20.2 versus 71.5 minutes (p=0.01), respectively. A slight decrease was seen in respiratory rate for the above 81 years cohort (16.1 versus 15.1 per minute, p = 0.04). Heart rate, total number of steps, and daily total active minutes did not change after TAVI compared to before TAVI for the overall group or for the different subgroups. There was no decrease in heart rate after TAVI despite a trend towards less use of beta blockers (pre 70% versus post 48% TAVI, p = 0.263). Univariate analysis of the Health Watch data could not identify a predictor for the good responders cohort (more than 40 meters of improvement during the 6MWT). Results are displayed in the supplemental material (Table S2).

**Table 3.** watch data pre versus post TAVI (overall cohort and good responders cohort)

| Parameter | Pre TAVI (n=97) | Post TAVI (n=75) | p-value | Good responders preTAVI (n=43) | p value* |
| --- | --- | --- | --- | --- | --- |
| Resting heart rate (1/min) | 62.5 ± 8.9 | 62.3 ± 8.2 | 0.88 | 62.6 ± 10.3 | 0.90 |
| Respiratory rate at rest (1/min) | 16.5 ± 1.9 | 16.2 ± 1.8 | 0.54 | 16.5 ± 2 | 0.70 |
| Heart rate (1/min) | 69.9 ± 8.3 | 69.5 ± 7.3 | 0.72 | 70.2 ± 9.6 | 0.95 |
| Heart rate during sleep (1/min) | 63.9 ± 8.9 | 63.3 ± 8.5 | 0.57 | 64.3 ± 10 | 0.61 |
| Respiratory rate during sleep (1/min) | 16.1 ± 2.1 | 15.9 ± 2.1 | 0.92 | 16.1 ± 2.3 | 0.67 |
| Daily percentage of HR observations < 60. bradycardia | 10.1 [1.2-34.5] | 14.7 [2.4-35.4] | 0.92 | 13.3 [0.8-3.0] | 0.46 |
| Daily percentage of HR observations > 100. tachycardia | 1.3 [0.3-2.9] | 1.3 [0.5-2.2] | 0.96 | 1 [0.3-1.0] | 0.86 |
| Daily total number of steps | 3586 [2607-4946] | 4341 [2093-6083] | 0.36 | 3633 [2763.0-5135.0] | 0.18 |
| Daily cumulative active energy expenditure (kcal) | 718.5 ± 206.5 | 722.6 ± 226.6 | 0.98 | 733 ± 221.4 | 0.15 |
| Daily cumulative total energy expenditure (kcal) | 2313.3 ± 401.3 | 2296.1 ± 436.6 | 0.72 | 2310.7 ± 428.1 | 0.19 |
| Slope of log(HR/TEE) | 0.27 ± 0.1 | 0.4 ± 1.0 | 0.26 | 0.26 ± 0.1 | 0.04 |
| Daily sleep time (hours) | 7.9 ± 1.8 | 10.1 ± 15.7 | 0.23 | 8.2 [7.0-8.7] | 0.28 |
| Daily basal activity time (min) | 209.0 [173 – 253] | 198.0 [167-262] | 0.43 | 209.0 [173.4-261.3] | 0.94 |
| Daily light activity time (min) | 183.1 ± 83.8 | 190.3 ± 100.3 | 0.63 | 195.2 ± 88.8 | 0.63 |
| Daily moderate activity time (min) | 48.8 ± 60.7 | 58.3 ± 51.3 | 0.10 | 20.2 [8.8-66.3] | 0.01 |
| Daily high activity time (min) | 0.0 [0.0-0.0] | 0.0 [0.0-1.5] | 0.18 | 0.0 [0.0-0.0] | 0.19 |
| Daily total active (min) | 101.9 ± 91.4 | 102.3 ± 71.9 | 0.98 | 108.3 ± 100.3 | 0.41 |

Abbreviations: HR = Heart Rate; TEE = Total Energy Expenditure; min = minutes. Summary values represent mean ± standard deviation or median with [interquartile ranges]. \*
p values good responders cohort preTAVI versus good responders post TAVI

### Energy Efficiency of the Cardiovascular system

The cardiac energy expenditure slope (CEES) – the slope of the fitted line in Figure 3 between heart rate (HR) and total energy expenditure (TEE) on a log scale – serves as an indicator for the energy efficiency of the cardiovascular system. CEES did not change significantly before versus after TAVI for the overall cohort (p value = 0.26), but for the good responders cohort there was a significant increase in CEES (p=0.04).

### 6MWT and Questionnaire (transformed WHOQOL-BREF)

The distance on the 6MWT increased after TAVI compared to before (342.8 versus 289.7 meters, p<0.001) both for the total cohort as well as for all subgroups. An improvement in the physical limitation score (domain 1 of the questionnaire) could be seen in the overall group (54.5 versus 61.4, p=0.005). In subgroup analyses, ‘male’ (55.4 versus 62.0, p=0.03), ‘below 81 years’ (51.8 versus 61.1, p=0.001) and ‘below 85 years’ (54.4 versus 60.8, p=0.013) similarly showed an improvement. Results from the other domains (psychological, level of independence, social relationships, overall) did not change for the total cohort. However, we detected an improvement in the psychological domain of the ‘above 81 years’ cohort (67.6 versus 71.7, p value = 0.025) and in the overall score of the ‘below 81 years’ cohort (248.4 versus 267.1, p value = 0.009). The 6MWT and quality of life questionnaire are summarized in Table 4 and Table 5, respectively.

**Table 4.**
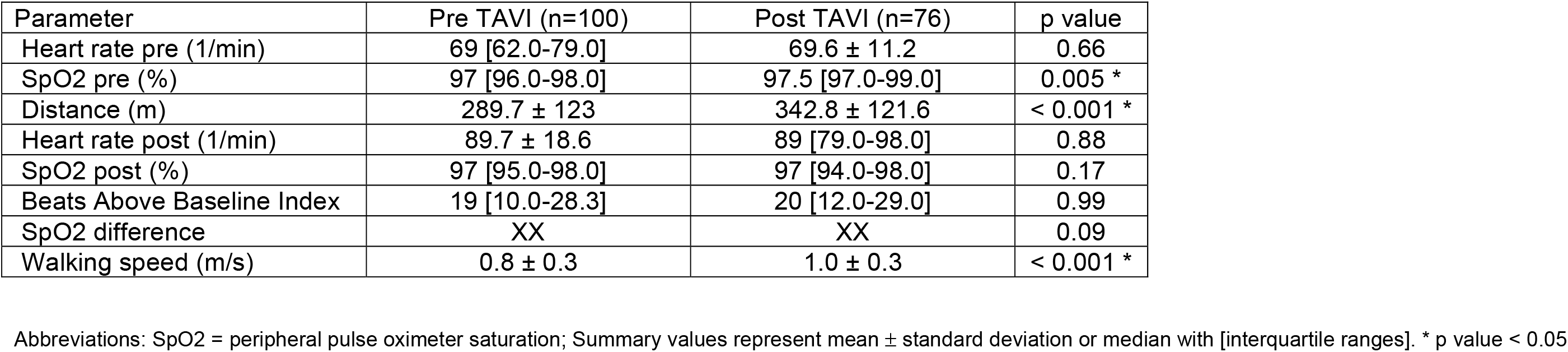
six minute walking test (6MWT) pre versus post TAVI

**Table 5.** Quality of life questionnaire pre versus post TAVI

|  | Pre TAVI<br>(n=100) | Post TAVI (n=76) | p value |
| --- | --- | --- | --- |
| Domain I - physical | 54.5 ± 18.7 | 61.4 ± 19.7 | 0.005* |
| Domain II - psychological | 68.1 ± 14.7 | 67.4 ± 14.4 | 0.64 |
| Domain III - Level of independence | 65.3 ± 15.5 | 69 [56.0-75.0] | 0.88 |
| Domain IV - social relationships | 71.4 ± 15.2 | 73.5 ± 15.8 | 0.31 |
| Overall score | 258.8 ± 53.6 | 268.7 ± 58.7 | 0.16 |

Abbreviations: Summary values represent mean ± standard deviation or median with [interquartile ranges]. \* p value < 0.05

## DISCUSSION

To our knowledge, this is the first study in which extensive one-week physiologic data before and after TAVI was assessed using a sophisticated wearable sensor, the Philips Health Watch. Watch parameters like activity time and step count did not increase after TAVI for the overall group, in contrast to significant improvements in 6MWT (53 meters, or 18% over baseline) and physical limitation score from the questionnaire (7 points, or 13% over baseline). The increase in 6MWT mirrors results obtained in a randomized trial: 254 meters at baseline, 288 meters at 30 days, and 297 meters 1 year after TAVI. ^12^

One explanation for our findings is that a relatively older population truly does not increase daily activity after TAVI, simply because they do not have to or do not want to (i.e. *lack of necessity or motivation)*. In such cases, step count and heart rate will not and do not have to increase. However, 6MWT increased significantly after 1 year in a cohort randomized to TAVI compared to no change in those randomized to medical therapy^13^, arguing against a Hawthorne effect. A second explanation for our findings is that quality of life questionnaires and 6MWT^4^ produce an *abundance of motivation* in the hospital setting that does not correspond to daily life. In this case the findings on standard tests (6MWT, QoL questionnaires) would appear improved, whereas parameters on the health watch would remain unchanged. Thus both tests may be correct given their circumstances. A third explanation is that the health watch parameters contain bias or imprecision that limits their paired comparison, even in a reasonably sized cohort such as our own. This seems to be an interesting topic for further research.

Novel metrics like the Cardiovascular Energy Expenditure Slope (CEES) may be valuable parameters in the future or in other settings for evaluating the energy efficiency of the heart. The steepness of the slope objectively quantifies the relationship between heart rate and total energy expenditure. A less steep slope corresponds with a lower CEES value. In such case more energy is needed to maintain heart rate than when the slope is steeper (and CEES value higher). There was a significant increase in CEES for the good responders cohort and this was also accompanied by a significant increase in moderate activity time. There was no significant improvement in CEES or moderate daily activity time in patients that had no or moderate (< 40 meters) improvement in the 6MWT after TAVI. This novel metric could be used in future research as a tool to identify patient improvement after TAVI intervention, independent of subjective variables. More research on this metric is warranted.

How to best identify patients whose symptoms benefit from treatment with TAVI remains an important and unanswered clinical question. A post-hoc analysis of a randomized trial of TAVI compared to a surgical valve procedure demonstrated that 36% of patients had no change in 6MWT after 30 days and 12 months, and 23-28% demonstrated no improvement on their QoL questionnaire scores (albeit using a different tool than in our cohort). ^12^ When considering an intervention, both procedural risks and economic costs should be balanced against the potential improvement in quality of life. Since the patients that commonly qualify for TAVI treatment are relatively older and more frail, an increase in physical performance can be equally or even more important than extending life expectancy. With increasing costs in health care, the benefit of an intervention should be clear and personalized^14^, and for TAVI this cost-benefit ratio has been disputed^15^. Unfortunately, the overall findings from this study cannot identify patients using the health watch who would be expected to have an above average response.

The impact of health watches and other sensor technologies on cardiologic care warrants more research. The Apple Heart study^7^ found an irregular heart rhythm in only 0.52% of over 400,000 people followed for 8 months, of whom just 21% completed further testing and of these a 34% minority were ultimately diagnosed with atrial fibrillation – a paltry yield of 153 people, or <<0.1% of the total. Findings such as these show the inevitable tradeoffs between mass testing versus pre-test probability of an actionable diagnosis.

### Limitations

Not all patients had available follow-up data, including 11 patients who died. However, follow-up and health watch data was complete in over 80% of patients. The second health watch measurement was performed 3 months after the procedure as that timing fit best with the local follow-up protocol. It might be speculated that not all patients have completely recovered at 3 months already. However, most studies comparing TAVI to surgical aortic valve replacement show a good functional improvement for the TAVI cohort at 30 days and 6 months^16^ compared to the SAVR group, while the TAVI cohort does not increase further towards 1 year. This implies that most patients are already at full capacity at 3 months follow-up.

## CONCLUSIONS

This is one of few studies before and after TAVI with extensive, one-week functional assessment with a sophisticated wearable sensor, the Philips Health Watch. Data from the health watch did not register an increase in activity time, total step count, or other parameters after TAVI, whereas traditional 6MWT and QoL assessment did improve. Watch-based parameters such as these might be less appropriate for measurement of treatment effect in the TAVI population. However our findings relating to the good responder subpopulation suggest that using data such as the CEES parameter derived from the data from a wearable device, might become useful to objectively identify patient improvement after TAVI intervention. This seems to be interesting for further research.

## Data Availability

The data that support the findings of this study are available from the corresponding author, RE, upon reasonable request.

## ABBREVIATIONS

6MWT: 6-minute walking test
AS: Aortic Stenosis
SAVR: Surgical Aortic Valve Replacement
TAVI: Transcatheter Aortic Valve Implantation
MET: Metabolic Equivalent Task
TEE: Total Energy Expenditure
CEES: Cardiovascular Energy Expenditure Slope
HR: Heart Rate
CO: Cardiac Output
SV: Stroke Volume
QoL: Quality of Life

## SUPPLEMENTAL MATERIAL

**Table S1.** Procedural data

| Characteristic | Total (n=97) | Male (n=56) | Female (n=41) | p value |
| --- | --- | --- | --- | --- |
| TAVI Device |  |  |  |  |
| Allegra | 2 (2%) | 1 (2%) | 1 (2%) |  |
| Edwards | 47 (48%) | 31 (55%) | 16 (39%) |  |
| Evolute | 48 (49%) | 24 (43%) | 24 (59%) |  |
| Post-operative |  |  |  |  |
| Procedural death | 1 (1%) | 0 (0%) | 1 (2%) | 0.42 |
| LBBB | 22 (23%) | 14 (25%) | 8 (20%) | 0.50 |
| Pacemaker implantation | 10 (10%) | 6 (11%) | 4 (10%) | 0.86 |
| Major bleeding | 15 (15%) | 10 (18%) | 5 (12%) | 0.45 |
| CVA | 1 (1%) | 0 | 1 (2%) | 0.43 |
| Coronary occlusion | 1 (1%) | 0 | 1 (2%) | 0.43 |

Abbreviations: TAVI = transcatheter aortic valve implantation; LBBB = left bundle branch block; CVA = cerebrovascular accident. Summary values represent number (%).

**Table S2.**
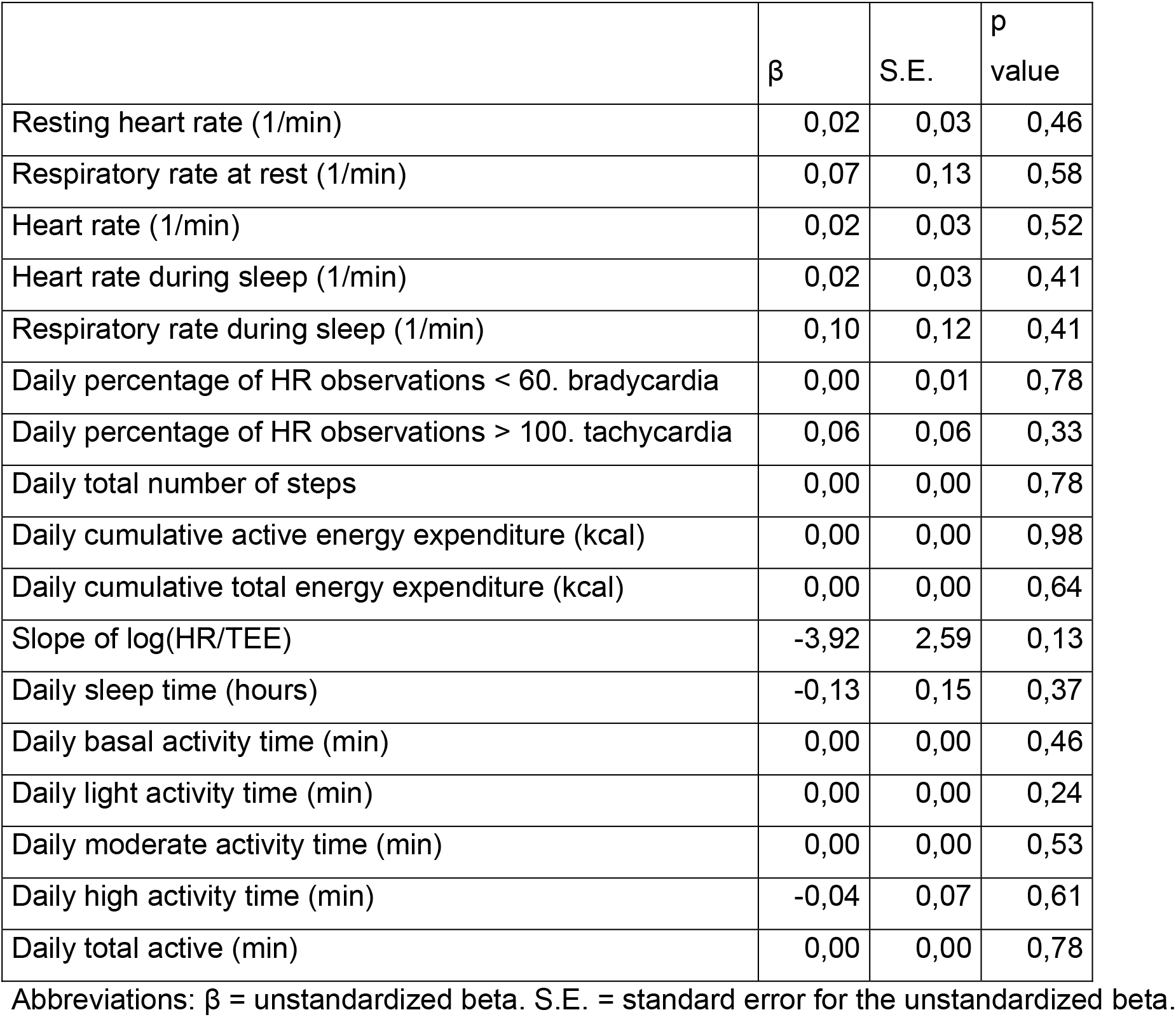
Univariate analysis of good responders watch data pre-TAVI

**Table S3.** Good responders watch data pre versus post TAVI

|  | Good responders<br>pre TAVI (n=43) | Good responders<br>post TAVI (n=40) | p value |
| --- | --- | --- | --- |
| Resting heart rate | 62.6 ± 10.3 | 62.3 ± 7.9 | 0.90 |
| Respiration rate at rest | 16.5 ± 2 | 16.2 ± 1.9 | 0.70 |
| Heart rate | 70.2 ± 9.6 | 70.2 ± 7 | 0.95 |
| Heart rate during sleep | 64.3 ± 10 | 63.4 ± 7.8 | 0.61 |
| Respiration rate during sleep | 16.1 ± 2.3 | 15.8 ± 2.3 | 0.67 |
| Daily percentage of HR<br>observations < 60, bradycardia | 13.3 [0.8-3.0] | 15.0 [3.5-35.5] | 0.46 |
| Daily percentage of HR<br>observations > 100, tachycardia | 1 [0.3-1.0] | 1.6 [0.7-2.7] | 0.86 |
| Daily total number of steps | 3633 [2763.0-5135.0] | 4488.5 [2965.5-6664.5] | 0.18 |
| Daily cumulative active energy<br>expenditure (kcal) | 733 ± 221.4 | 790.6 ± 218.4 | 0.15 |
| Daily cumulative total energy<br>expenditure (kcal) | 2310.7 ± 428.1 | 2372.8 ± 466.1 | 0.19 |
| Slope of log(HR/TEE) | 0.26 ± 0.1 | 0.3 ± 0.1 | 0.04 |
| Daily sleep time (hrs) | 8.2 [7.0-8.7] | 8.3 [7.3-9.2] | 0.28 |
| Daily basal activity time (mns) | 209.0 [173.4-261.3] | 200.1 [166.2-264.6] | 0.94 |
| Daily light activity time (mns) | 195.2 ± 88.8 | 204.9 ± 77.1 | 0.63 |
| Daily moderate activity time (mns) | 20.2 [8.8-66.3] | 66.8 [29.5-113.4] | 0.01 |
| Daily high activity time (mns) | 0.0 [0.0-0.0] | 0 [0.0-2.0] | 0.19 |

